# ReIGNITE RT Boost: An international study testing the accuracy and feasibility of using restriction spectrum imaging (RSI) MRI to guide intraprostatic tumor target volume for radiotherapy boost

**DOI:** 10.1101/2022.12.13.22283420

**Authors:** Asona J. Lui, Karoline Kallis, Allison Y. Zhong, Troy S. Hussain, Christopher Conlin, Leonardino A. Digma, Nikki Phan, Ian T. Mathews, Deondre D. Do, Mariluz Rojo, Roshan Karunamuni, Joshua Kuperman, Anders M. Dale, Rebecca Rakow-Penner, Michael E. Hahn, Tyler M. Seibert

## Abstract

In a phase III randomized trial, adding a radiation boost to visible tumor(s) on MRI improved prostate cancer disease-free and metastasis-free survival without additional toxicity. However, radiation oncologists’ ability to identify prostate tumors is critical and represents a major barrier to widely adopting intraprostatic tumor radiotherapy boost for patients. We previously developed a quantitative diffusion MRI biomarker for prostate cancer, called the Restriction Spectrum Imaging restriction score (RSIrs), that has been shown to improve radiologist identification of clinically significant prostate cancer.

42 radiation oncologists (participants) from multiple, international institutions contoured prostate tumors on 40 patient cases using standard MRI with or without RSIrs map, producing 1646 target volumes. Use of RSIrs maps significantly improved all evaluated accuracy metrics, including participants’ percent overlap with consensus expert target volume (73% vs. 42%, p<0.001). A mixed effects model confirmed that RSIrs maps were the main variable driving the improvement in all metrics. System Usability Scores indicated RSIrs maps significantly improved the contouring experience (72 vs. 58, p<0.002). The expert-defined tumor was completely missed 158 times on standard MRI alone and only 19 times with RSIrs maps. RSIrs maps improve the accuracy of target delineation for prostate tumor boost.

**Patient Summary:** Adding an extra boost of radiation to tumor(s) visible on MRI has been shown to prevent cancer recurrence and cancer spread beyond the prostate without adding additional side effects; however, drawing the prostate tumor on MRI is difficult, and most radiation oncologists have not been trained to do this. We have developed an advanced MRI technique (RSIrs maps) that increases tumor visibility. We found that RSIrs maps improve radiation oncologists’ accuracy in targeting prostate tumors.

## Manuscript

Standard radiation therapy for aggressive prostate cancer (PCa) treats the entire prostate gland with an equally distributed radiation dose of 70-80 Gy. In a recent phase III randomized controlled trial, addition of a focal radiotherapy boost to PCa lesions visible on MRI (hereafter called tumors) up to 95 Gy increased disease-free survival from 86% to 93% at 7 years when compared to standard dose delivery.^1^ Both local control and regional/distant metastasis-free survival were also improved.^2^ Remarkably, these patient benefits did not come at the cost of additional short- or long-term toxicity.^2,3^ However, radiation oncologists’ ability to identify prostate tumors on MRI is critical to harnessing the benefits of radiotherapy boost for patients.

In the FLAME trial, expert radiologists assisted with target identification, but even subspecialty-trained, experienced radiologists show substantial variability in lesion identification.^4,5^ Tumor identification has not been part of most radiation oncology training and presents a major barrier to widely adopting intraprostatic tumor boost for prostate radiotherapy.

We previously developed a quantitative diffusion MRI biomarker for PCa, called the Restriction Spectrum Imaging restriction score (RSIrs), that significantly improves diagnostic utility over conventional MRI.^6,7^ RSI is a sophisticated but practical approach that models tissue microcompartments to highlight the restricted intracellular diffusion characteristic of higher-grade PCa.^8^ We hypothesized that using RSIrs maps would improve radiation oncologists’ accuracy for PCa tumor delineation.

42 radiation oncologist participants with varied levels of experience were enrolled as participants in our study **(Supplemental Table 1**). Participants still in training were required to have previously completed a prostate cancer radiation oncology clinical rotation. All study recruitment materials, communications, and procedures were approved by the UC San Diego Institutional Review Board (IRB).

Study participants were asked to contour tumors on 20 patient cases—half with conventional MRI alone and half with conventional MRI plus RSIrs—in each of two sessions at least 1 month apart. Without informing the participants, 10 of the cases from the first session were interspersed within the second session but with RSIrs either removed or added from the case. Conventional MRI included *T*_2_-weighted, ADC, and DWI (b=0 and b=2000 s/mm^2^). RSIrs maps were displayed as an overlay on the anatomic *T_2_*-weighted images **(Supplemental Figure 1)**. Participants contoured tumor volumes using the MIM Zero Footprint™ (ZFP) platform. They were also provided with basic clinical information for each case: patient age, PSA at time of MRI, Gleason score, number and location of positive cores, and the radiologist’s description of the location and size of the lesion. Expert volumes were created by consensus interpretation by two board-certified, sub-specialist GU radiologists, R.R.P. (5 years’ experience) and M.E.H. (7 years’ experience), using conventional multiparametric MRI (including dynamic contrast enhanced images) and the clinical/pathologic information for each case. They were assisted by a radiation oncologist, A.J.L. (3 years’ experience). All volumes were exported as binary masks and analyzed in MATLAB R2021b (Mathworks, Natick, MA).

1646 participant volumes were compared to the relevant consensus expert volumes using four metrics of accuracy: percent overlap with expert volume, Dice coefficient, conformal number, and maximum distance beyond expert volume **(Table 1 and Supplemental Figure 2)**. The addition of RSIrs maps to conventional MRI significantly improved all accuracy metrics by at least 50%, including overlap with expert volume (37.8% vs. 73.2%, p<0.0001) **(Table 1)**. There were 158 complete misses with conventional MRI alone and only 19 complete misses with addition of RSIrs maps. Improvement in accuracy and reduction of variability are illustrated in **Figure 1 and Supplemental Figure 1** for representative cases.

**Table 1:**
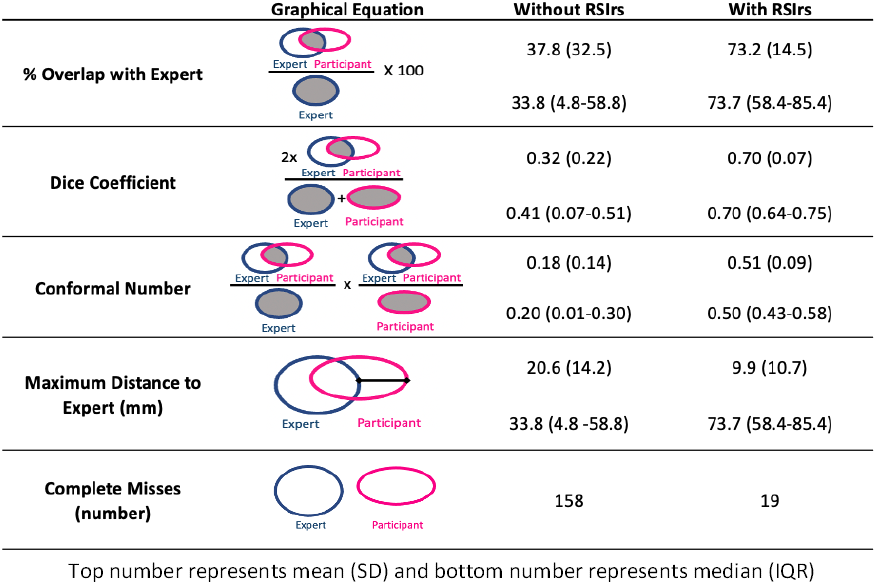
Summary accuracy statistics comparing participant to expert.

**Figure 1:**
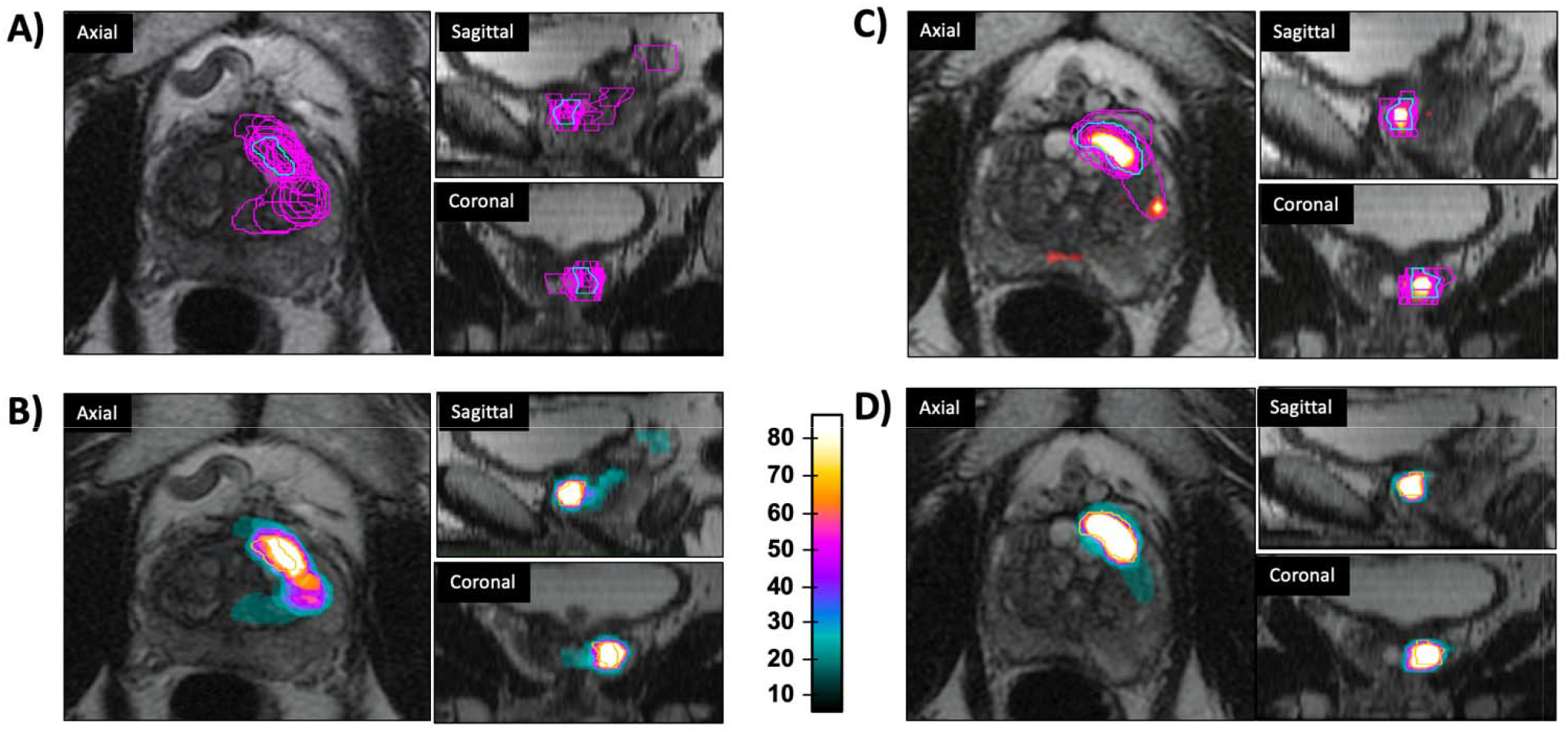
RSIrs maps improve the accuracy of participant target volume. **(A and C)** A representative selection of participant volumes is overlaid with the expert. **A)** with conventional MRI or **C)** with conventional MRI with RSIrs map (orange heatmap). **(B and D)** Heatmap displaying the variation of all participant volumes on a single case. Each color represents the given frequency of inclusion of that voxel in participant volumes and the expert is now highlighted in orange. **B)** with standard MRI or **D)** with standard MRI with RSIrs map.

Mixed-effects models demonstrated that use of RSIrs maps was the main driver of improvement in accuracy metrics in our study, not participants’ experience with prostate tumor boost **(Supplemental Tables 2-5)**. Use of RSIrs maps was independently associated with improvement in all four accuracy metrics and was the only independent predictor of accuracy for three metrics (percent overlap, Dice coefficient, and conformal number).

After completing the volumes, participants were given a System Usability Scale (SUS) questionnaire adapted from the US Department of Health and Human Services.^9^ Responses to the SUS questionnaire (on a scale of 0 to 100, with >70 considered “acceptable” and ≤50 considered “poor” usability) were analyzed in the standard manner, interpreted to evaluate the relative difficulty and preference for contouring with and without RSIrs maps. SUS score improved significantly with the use of RSIrs maps (57 vs. 72, p<0.001). Notably, target delineation on conventional MRI was rated poorly, with 36% of participants reporting an SUS score≤50 and only 26% reporting SUS score >70. With the addition of RSIrs maps, the proportion of participants reporting a “poor” experience was reduced to 5% and the proportion reporting “acceptable” experience increased to 48% **(Supplemental Figure 3)**. This result underscores the inherently challenging nature of tumor contouring and highlights the need for new methods and training so that tumor radiotherapy boost can become standard for patients with PCa.

Limitations of this study include the use of images from a single scanner and a single institution. Ongoing studies are evaluating the quantitative reproducibility of RSIrs maps across scanner platforms. This study used expert radiologist consensus volumes as the reference, in conjunction with the clinical and pathological (biopsy) information. We have previously demonstrated improved voxel-wise PCa detection with RSIrs^6^ and are currently collecting data to compare RSIrs to whole-mount histopathology to evaluate tumor extent. These efforts might yield improved accuracy even beyond the radiologist standard used in the present study and in the FLAME trial that established the benefit of focal tumor boost.

We report that RSIrs maps significantly improve the accuracy and reduce the variability of target delineation for prostate tumor radiotherapy boost. RSIrs maps can be generated from brief acquisitions on standard MRI scanners—approximately two minutes of additional scan time for the maps in this study. An implementation of RSI software is cleared by the FDA for prostate MRI and already commercially available on multiple scanner/vendor platforms. RSIrs maps have the potential to increase the feasibility of widely implementing the benefits of focal tumor boost for patients undergoing prostate radiotherapy.

## Take Home Message

RSIrs maps improve radiation oncologists’ accuracy for target delineation for prostate tumor radiotherapy boost. RSIrs maps have the potential to increase the feasibility of widely implementing the benefits of focal tumor boost for patients undergoing prostate radiotherapy.

## Data Availability

De-identified data are available to bona fide researchers for non-commercial use upon request.

## Data Availability Statement

De-identified data are available to bona fide researchers for non-commercial use upon request.

---

**Supplemental Table 1:**
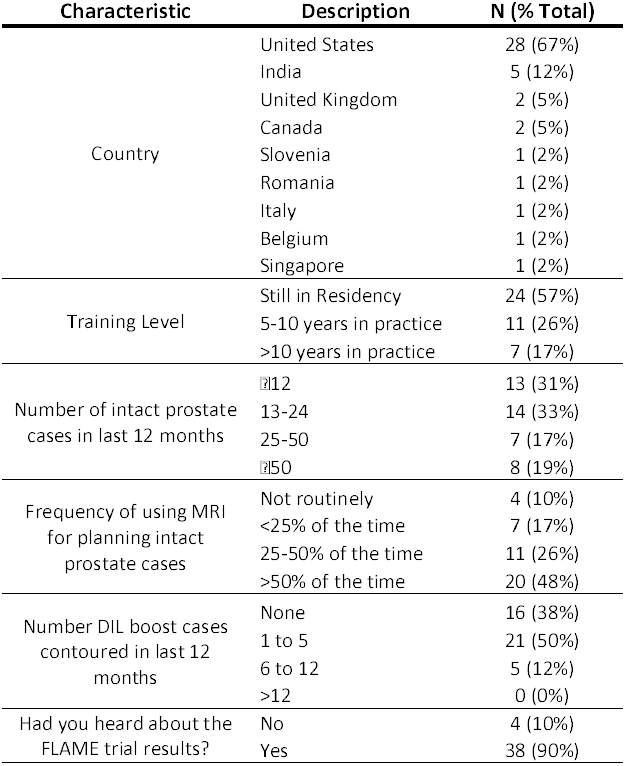
Participant Characteristics.

**Supplemental Table 2:**
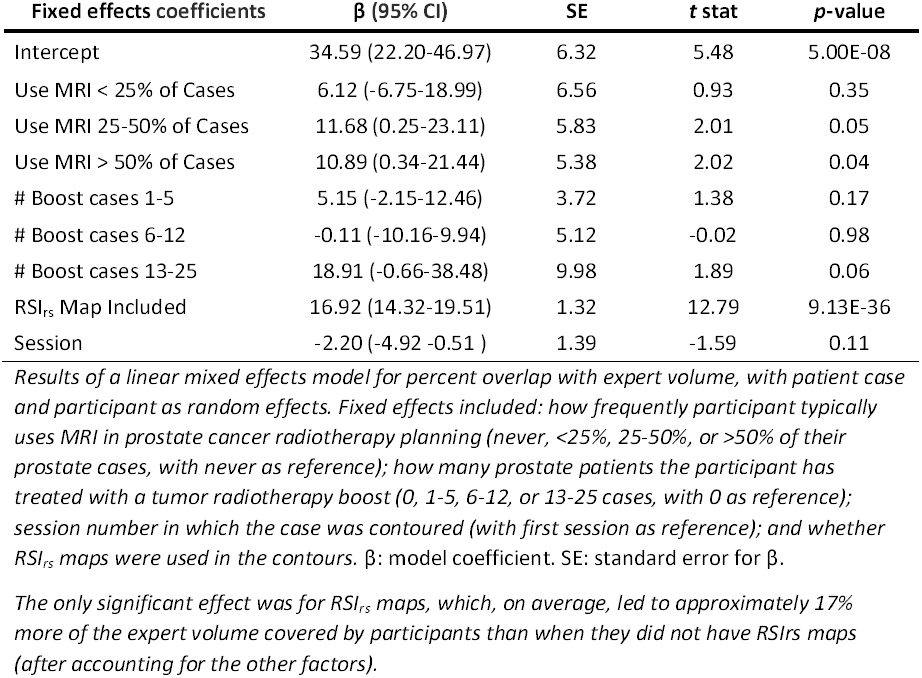
Linear mixed effects model for Percent Overlap with Expert Volume.

| Fixed effects coefficients | $\beta$ (95% CI) | SE | t stat | p-value |
| --- | --- | --- | --- | --- |
| Intercept | 34.59 (22.20-46.97) | 6.32 | 5.48 | 5.00E-08 |
| Use MRI < 25% of Cases | 6.12 (-6.75-18.99) | 6.56 | 0.93 | 0.35 |
| Use MRI 25-50% of Cases | 11.68 (0.25-23.11) | 5.83 | 2.01 | 0.05 |
| Use MRI > 50% of Cases | 10.89 (0.34-21.44) | 5.38 | 2.02 | 0.04 |
| # Boost cases 1-5 | 5.15 (-2.15-12.46) | 3.72 | 1.38 | 0.17 |
| # Boost cases 6-12 | -0.11 (-10.16-9.94) | 5.12 | -0.02 | 0.98 |
| # Boost cases 13-25 | 18.91 (-0.66-38.48) | 9.98 | 1.89 | 0.06 |
| RSI<sub>rs</sub> Map Included | 16.92 (14.32-19.51) | 1.32 | 12.79 | 9.13E-36 |
| Session | -2.20 (-4.92 -0.51 ) | 1.39 | -1.59 | 0.11 |

*Results of a linear mixed effects model for percent overlap with expert volume, with patient case and participant as random effects. Fixed effects included: how frequently participant typically uses MRI in prostate cancer radiotherapy planning (never, <25%, 25-50%, or >50% of their prostate cases, with never as reference); how many prostate patients the participant has treated with a tumor radiotherapy boost (0, 1-5, 6-12, or 13-25 cases, with 0 as reference); session number in which the case was contoured (with first session as reference); and whether RSI<sub>rs</sub> maps were used in the contours. $\beta$ : model coefficient. SE: standard error for $\beta$ .*
*The only significant effect was for RSI<sub>rs</sub> maps, which, on average, led to approximately 17% more of the expert volume covered by participants than when they did not have RSI<sub>rs</sub> maps (after accounting for the other factors).*

**Supplemental Table 3:**
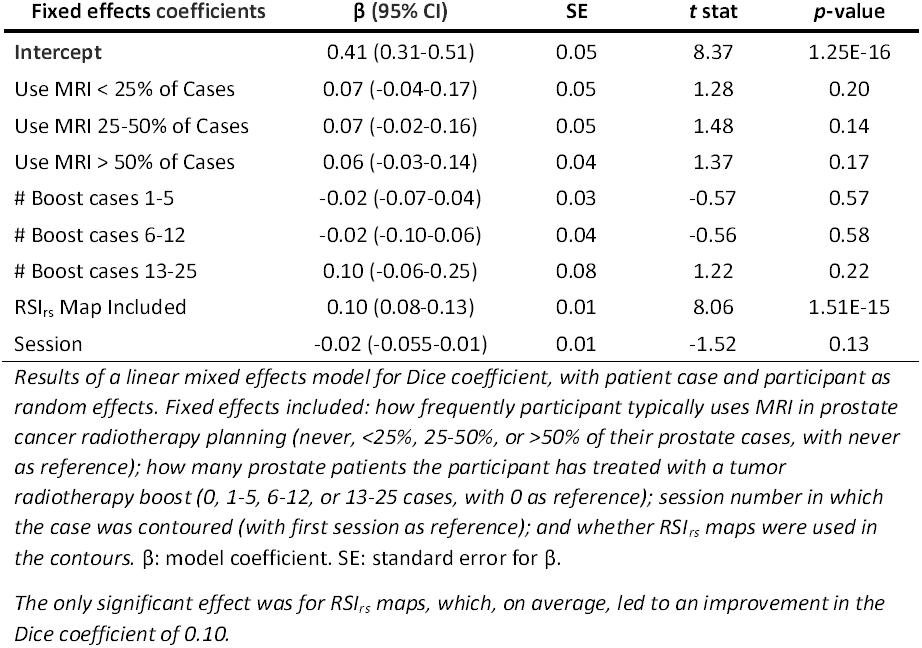
Linear mixed effects model for Dice Coefficient.

| **Fixed effects coefficients** | **$\beta$ (95% CI)** | **SE** | **t stat** | **p-value** |
| --- | --- | --- | --- | --- |
| **Intercept** | 0.41 (0.31-0.51) | 0.05 | 8.37 | 1.25E-16 |
| Use MRI < 25% of Cases | 0.07 (-0.04-0.17) | 0.05 | 1.28 | 0.20 |
| Use MRI 25-50% of Cases | 0.07 (-0.02-0.16) | 0.05 | 1.48 | 0.14 |
| Use MRI > 50% of Cases | 0.06 (-0.03-0.14) | 0.04 | 1.37 | 0.17 |
| # Boost cases 1-5 | -0.02 (-0.07-0.04) | 0.03 | -0.57 | 0.57 |
| # Boost cases 6-12 | -0.02 (-0.10-0.06) | 0.04 | -0.56 | 0.58 |
| # Boost cases 13-25 | 0.10 (-0.06-0.25) | 0.08 | 1.22 | 0.22 |
| RSI<sub>rs</sub> Map Included | 0.10 (0.08-0.13) | 0.01 | 8.06 | 1.51E-15 |
| Session | -0.02 (-0.055-0.01) | 0.01 | -1.52 | 0.13 |

*Results of a linear mixed effects model for Dice coefficient, with patient case and participant as random effects. Fixed effects included: how frequently participant typically uses MRI in prostate cancer radiotherapy planning (never, <25%, 25-50%, or >50% of their prostate cases, with never as reference); how many prostate patients the participant has treated with a tumor radiotherapy boost (0, 1-5, 6-12, or 13-25 cases, with 0 as reference); session number in which the case was contoured (with first session as reference); and whether RSI<sub>rs</sub> maps were used in the contours. $\beta$ : model coefficient. SE: standard error for $\beta$ .*
*The only significant effect was for RSI<sub>rs</sub> maps, which, on average, led to an improvement in the Dice coefficient of 0.10.*

**Supplemental Table 4:** Linear mixed effects model for Conformal Number.

| Fixed effects coefficients | $\beta$ (95% CI) | SE | t stat | p-value |
| --- | --- | --- | --- | --- |
| Intercept | 0.25667 (0.18-0.34) | 0.04 | 6.46 | 1.38E-10 |
| Use MRI < 25% of Cases | 0.062082 (-0.02-0.15) | 0.04 | 1.48 | 0.14 |
| Use MRI 25-50% of Cases | 0.053633 (-0.02-0.13) | 0.04 | 1.44 | 0.15 |
| Use MRI > 50% of Cases | 0.050366 (-0.02-0.12) | 0.03 | 1.46 | 0.14 |
| # Boost cases 1-5 | -0.0098308 (-0.06-0.04) | 0.02 | -0.41 | 0.68 |
| # Boost cases 6-12 | -0.014149 (-0.08-0.05) | 0.03 | -0.43 | 0.67 |
| # Boost cases 13-25 | 0.07996 (-0.05-0.21) | 0.06 | 1.26 | 0.21 |
| RSI<sub>rs</sub> Map Included | 0.096686 (0.08-0.12) | 0.01 | 9.23 | 7.97E-20 |
| Session | -0.012154 (-0.03-0.01) | 0.01 | -1.11 | 0.27 |

*Results of a linear mixed effects model for conformal number, with patient case and participant as random effects. Fixed effects included: how frequently participant typically uses MRI in prostate cancer radiotherapy planning (never, <25%, 25-50%, or >50% of their prostate cases, with never as reference); how many prostate patients the participant has treated with a tumor radiotherapy boost (0, 1-5, 6-12, or 13-25 cases, with 0 as reference); session number in which the case was contoured (with first session as reference); and whether RSI<sub>rs</sub> maps were used in the contours. $\beta$ : model coefficient. SE: standard error for $\beta$ .*
*The only significant effect was for RSI<sub>rs</sub> maps, which, on average, led to an improvement in the conformal number of 0.10.*

**Supplemental Table 5:**
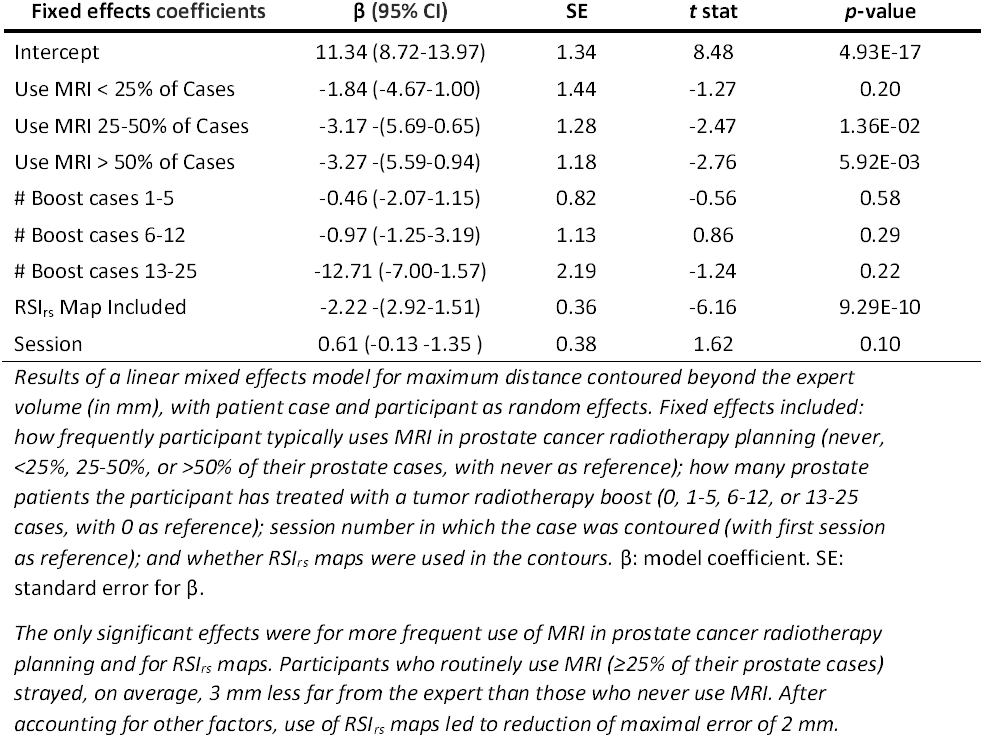
Linear mixed effects model for Maximum Distance beyond Expert Volume.

**Supplemental Figure 1:**
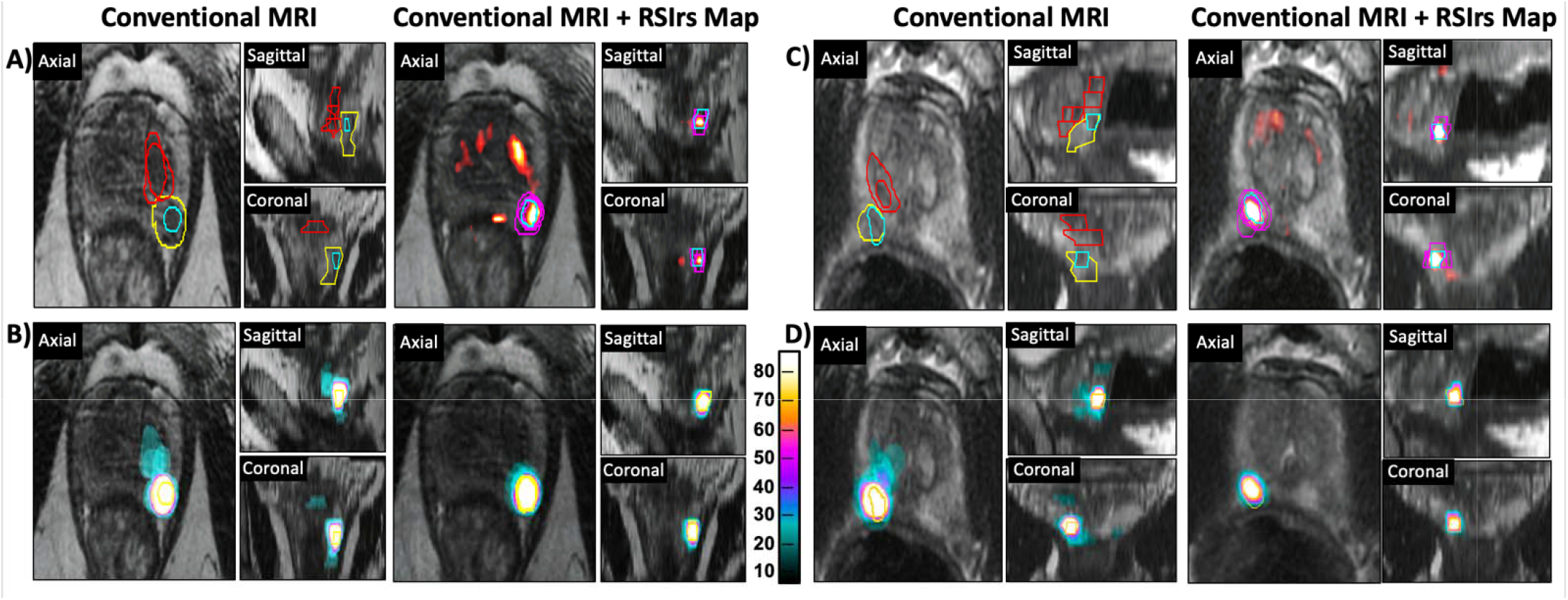
RSIrs maps improve the accuracy of participant target volumes. Example target volumes from two patient cases **(A/B and C/D)**where participants were provided Conventional MRI (T2-weighted, DWI, ADC)*(left)* images or conventional MRI with RSIrs map (orange heatmap)*(right)*. (A and C) Expert volumes are highlighted in Blue and participant volumes participant volumes highlighted by category relative to the expert target: Red = Complete Miss, Yellow= large distance to expert, Pink = Overlapping. **(B and D)** Heatmapi displaying the variation of all participant volumes on a single case. Each color represents the given frequency of inclusion of that voxel in participant volumes and the expert is now highlighted in orange.

**Supplemental Figure 2:**
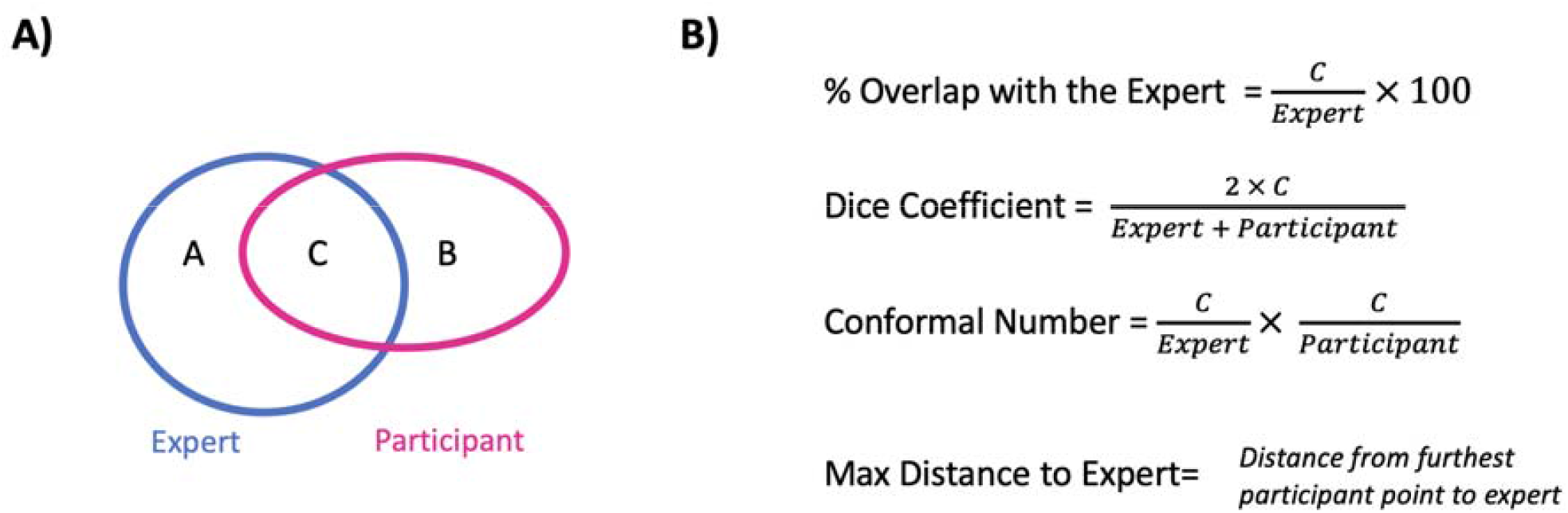
Accuracy metrics. **A)** Diagram representing overlapping “C” and non-overlapping “A” or “B” regions of expert and participant volumes used to calculate accuracy metrics. **B)** Equations for measures of accuracy used in data analysis.

**Supplemental Figure 3:**
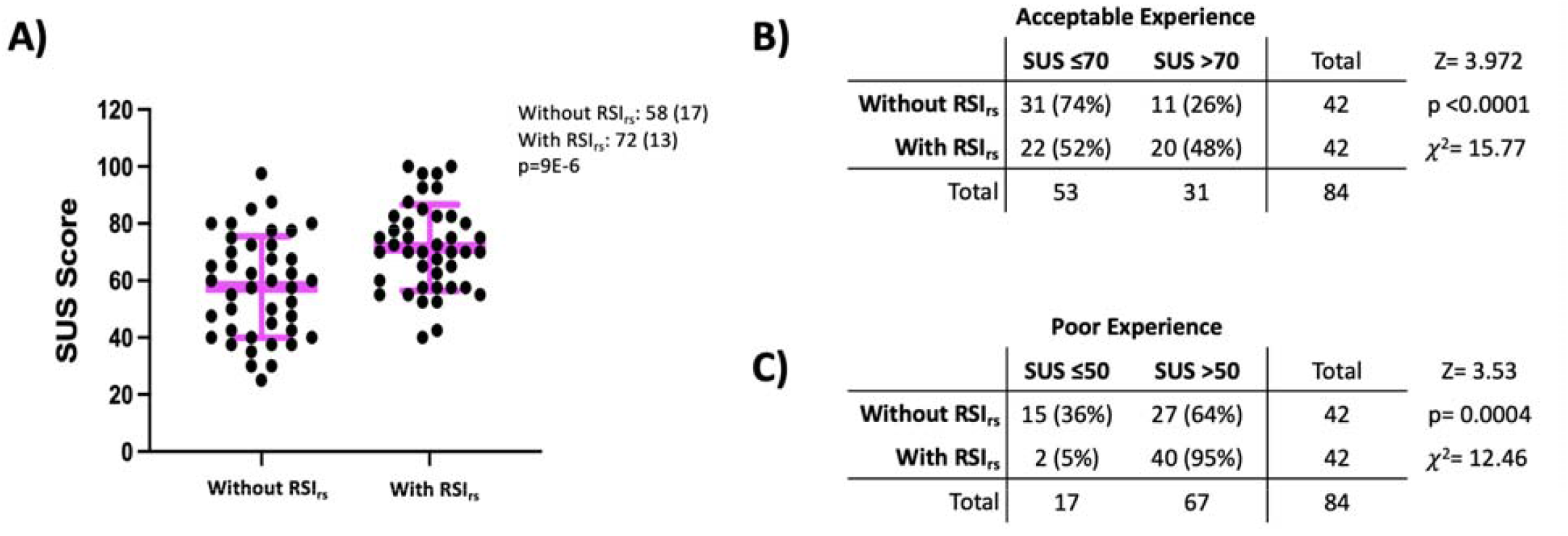
Summary of System Usability Survey results. **A)** Distribution of normalized SUS scores for all participants where figure legend and pink bars highlight Mean (SD). p-value is the result of a two-sided paired t-test. **(B and C)** 2x2 analysis of number of participants rating the contouring experience **B)** Acceptable (SUS > 70) and C) Poor (SUS ≤ 50). Percentages are percent of responses rating the contouring experience with RSI_rs_ (N=42) or Without RSI_rs_ (N=42).

